# Floor Sitting Rising in Adults with and without Disability: A Scoping Review Protocol

**DOI:** 10.1101/2024.03.20.24303720

**Authors:** Jehan Abdulmohsen Alomar, Haoyu Li, Keri Fisher, Miriam King, Lori Quinn

## Abstract

**Background:** Getting down to the floor and rising to a standing position (Floor Sitting-Rising | FSR) is a fundamental task for independent living and participation across the lifespan. Multiple studies have reported activity limitations in FSR among adults and individuals with musculoskeletal and neurological impairments. However, few studies have investigated FSR assessments and the body structures and functions contributing to FSR performance.

**Objectives:** To describe assessments that measure FSR in adulthood, including their psychometric properties, and to determine if impairments in body structures and functions contribute to limitations in FSR performance in adults with orthopedic or neurological disability.

**Design:** Google Scholar, Pubmed, CINHAL (Medline), and Rehab Measures will be searched for studies that report the full FSR transition. Studies must be original research in the adult population.

**Result:** We will categorize studies based on aims, study type, population characteristics, and abilities. We will narratively synthesize results, discuss potential personal and environmental factors influencing FSR, and identify the gaps in the literature to inform future research directions.

**Conclusion:** This review of FSR assessments will provide recommendations for methods to evaluate FSR and its movement strategies and consider impairments that may influence performance.

## Introduction

Getting down to the floor and rising to a standing position (Floor Sitting-Rising | FSR) is a fundamental task for independent living. (Fleming et al., 2008; Mulholland & Wyss, 2001) Socio-cultural traditions, societal norms, individual values, and the physical environment dictate daily activities (household, leisure, and occupational) that require deep squatting, kneeling, floor sitting, and rising from the floor. (Hewes, 1957; Lyman et al., 2019; Mohamed et al., 2015; Mulholland & Wyss, 2001) In eastern countries, such as Japan, China, and Saudi Arabia, FSR transition is frequently performed across the lifespan.(Acker et al., 2011; Fukuichi & Sugamura, 2022; Mohamed et al., 2015) In western countries, while children perform FSR transitions frequently, adults, particularly older adults, perform this task less frequently than in other countries. (Weingarten & Kaplan, 2015) This may contribute to the limitations in performing floor activities and the ability to return to standing after a fall later in life. (Ardali et al., 2019)

The FSR task has been used in rehabilitation research to investigate impairments, activity limitations, and participatory restrictions. Ferraz and colleagues used Timed FSR to assess for lower limb power among older adults with neurological impairments. (Ferraz et al., 2018). Gurley and colleagues highlighted that one-third of individuals discovered incapacitated or deceased at home on the floor were unable to independently rise to stand after a fall. (Gurley et al., 1996) Beyond its role in post-fall recovery, the FSR task serves as a foundational element for diverse participatory activities. Borchers and colleagues incorporated FSR into a comprehensive battery of tests to stratify participants with a neurological condition into community exercise classes. (Borchers et al., 2019). This underscores the importance of FSR in screening for impairments, activity limitations, and participatory restriction.

Several researchers have developed methods for evaluating both supine-to-stand and FSR. More than thirty studies used supine-to-stand to test movement time, report on movement strategies, or investigate the physiological changes in blood pressure in response to posture change. In a systematic review conducted in 2020, Cattuzzo and colleagues found inconsistencies in the methods of evaluating supine-to-stand. As a result, the group created a standardized supine-to-stand test protocol to be implemented. (Cattuzzo et al., 2020) Although some previous studies have examined the complete FSR task (Ardali et al., 2019; Brito et al., 2014), a review of the measurement properties of assessments encompassing the entire transitional sequence is currently lacking.

Given the heightened biomechanical complexity inherent in the entire FSR compared to supine to stand, a comprehensive analysis of the complete FSR task becomes especially valuable for discerning underlying motor impairments (Quinn et al., 2021) For example, Nagrajan and colleagues who used a newly developed FSR movement analysis tool to identify alterations in movement sequencing among older adults compared to their younger counterparts (Nagrajan & D’Souza, 2017). Notably, under dual-task conditions, the young adult group exhibited changes in their movement sequence, a phenomenon not observed in the older adult group. This observation suggests a potential role of anticipatory movement control processing in motor planning and adapting to task demands. Similar to the investigation by Nagrajan and colleagues, systematic observation of the FSR movement enables the formulation of theories regarding the underlying reasons for specific movement strategies. This, in turn, facilitates the diagnosis of movement impairments, driving evidence-based treatment decisions.

Individuals with orthopedic and neurological impairments have consistently worse performance on FSR assessments compared to their age-matched counterparts or reference datasets.(Alomar et al., 2020; Araújo et al., 2020; Ng et al., 2016) Furthermore, investigations into the connection of impairments to FSR performance have frequently shown associations between body structure integrity and limitations in FSR task performance. (Alomar et al., 2020; Ng et al., 2016) Within the rehabilitation community, a comprehensive review is currently absent regarding the influence of impairments on FSR movement performance. A synthesis of findings from studies encompassing diverse clinical populations holds the potential to unveil key movement strategies associated with specific demographics and impairments. This information will provide guidance to rehabilitation specialists, offering insights on what to observe and assess during clinical FSR assessment.

The aims of this scoping review are to (1) describe assessments that measure FSR, including their psychometric properties, and (2) to determine which body structures and functions contribute to FSR performance in adult populations. Our research follows an exploratory and descriptive approach, aiming to provide a comprehensive overview of existing evidence regarding FSR, including people with and without orthopedic or neurological impairments. This design allows us to establish inclusion criteria compatible with our broad aims and incorporate a wide range of quantitative study designs, offering the flexibility to adjust selection as needed while maintaining methodological rigor. We anticipate methodological heterogeneity among the studies, precluding us from conducting a quantitative synthesis (meta-analysis) of the data and deterring us from choosing a systematic review design. Similar to systematic reviews, a scoping review design will enable us to present a qualitative overview of the results, categorizing and extracting clinically relevant insights, identifying gaps, and providing recommendations for future research.

## Method

This protocol was drafted using the Preferred Reporting Items for Systematic Reviews and Meta-analysis Protocols (PRISMA). The aims and eligibility criteria were developed according to the PCC framework (participant, concept, and context).

Regarding the participants, we will include studies on adults with or without disabilities. The concept under study in this review is evidence mapping related to assessment measurement properties of the FSR task and the relationship between performance and impairments. We will consider all contexts and environments except FSR transition with assistance or from a wheelchair.

### Eligibility Criteria

The eligibility criteria for this review are as follows: (1) studies reporting the full transition from upright standing to sitting on the floor and returning to standing; (2) studies describing movement strategies and evaluating FSR among adults with or without musculoskeletal or neurological Impairments; (3) studies must be original research; We will exclude protocols, trial registration, and any resources examining the following: (1) one direction of the transfer; and (2) floor transfer where participants require external assistance and use of a wheelchair.

### Information resources

To identify potentially relevant documents, the following bibliographic databases were searched: Google Scholar, Pubmed, CINHAL (Medline), and Rehab Measure before November 2023. The search strategies were drafted with the assistance of an experienced librarian [Alan Foresta, Informationist at Teachers College, Columbia University] and further refined through team discussion. The final search strategy for all databases can be found in Table 1. The final search results were exported into Covidence v.2603 (Covidence.org, Melbourne, Australia), an online tool that facilitates and manages the collaborative process of systematic reviews. Duplicates were removed by study lead using Covidence v.2603.

**Table 1:**
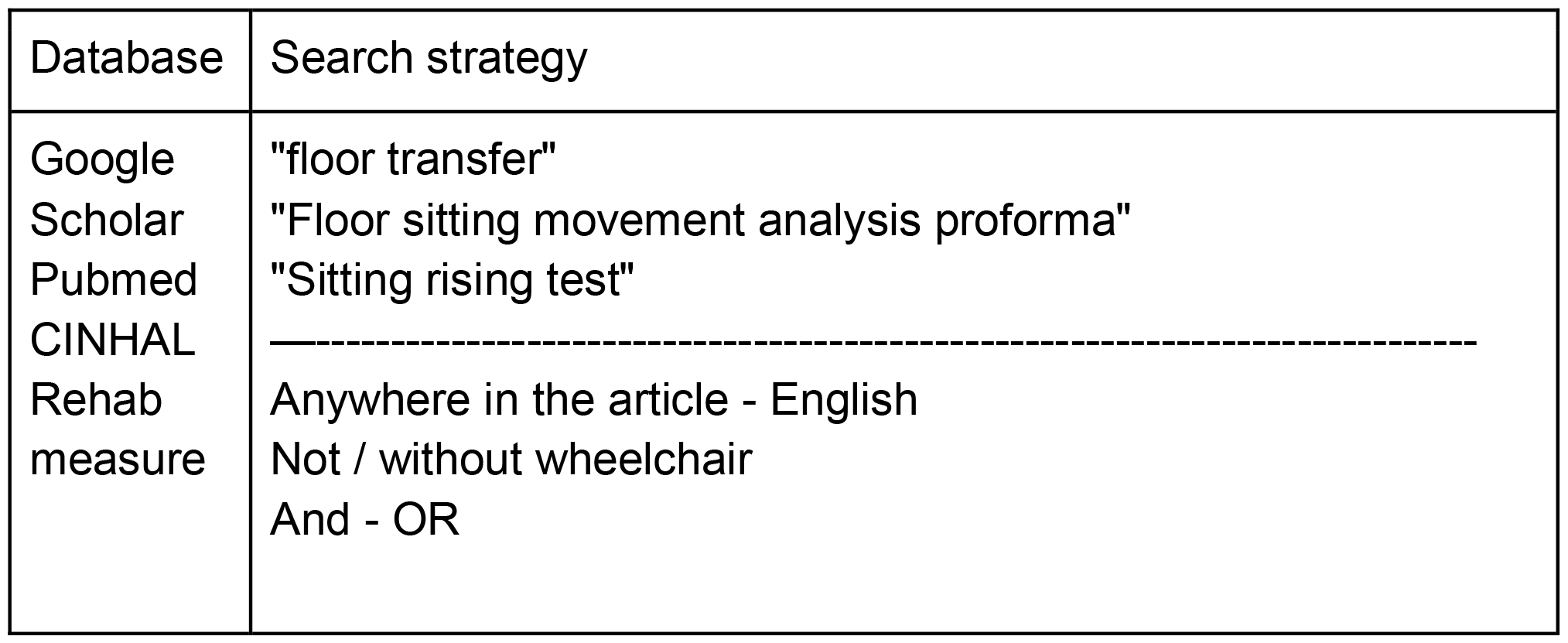
Search StrategyTable.

### Search

The search strategy will be modified for each database. We will consider FSR synonyms used in rehabilitation practice and research-controlled MeSH terms. We will use Boolean Operations (AND, OR, NOT) suitable for each database. FSR-specific search keywords are listed in Table 1. For example, 1320 documents were found using our search strategy for Google Scholar:

*“floor transfer” OR “Floor sitting movement analysis proforma” OR “Sitting rising test”*

### Selection of sources of evidence

Two reviewers will complete all the screening steps, and a third reviewer will consult to resolve disagreements. Three reviewers will screen the same 10 publications to increase consistency among reviewers and discuss their selection before beginning screening for this review.

## Data Charting

The study lead developed a data-charting form and jointly tested it with two other reviewers to determine if modifications were needed. The three reviewers will independently chart the data from 5 studies, discuss the results, and continuously update the data-charting form in an iterative process.

For full-text review, 4 reviewers working in pairs will sequentially extract from the full-text publications. Each study was verified by the study lead. Weekly meetings will be used to update the data extraction sheet and resolve disagreements on study extraction by consensus and discussion with other reviewers.

We will extract data on article characteristics (e.g, authors, journal, and year), the population (e.g., age and diagnosis), method (study design, aim, outcome), outcome (FSR assessment), and psychometrics (e.g., validity and reliability). Table 2 summarizes the relevant data included on the data charting spreadsheet.

**Table 2:** Data to be extracted.

|  |  |
| --- | --- |
| Publication Summary | Author, affiliation, country of affiliation, title, year of publication, journal, inclusion criteria. |
| Population | Total sample size, age, gender, race, ethnicity, diagnosis, time since diagnosis |
| Method | The type of research (e.g., intervention, qualitative, review, case study), setting, study aims or purpose, FSR definition, data collection method, and type of analysis. |
| Outcomes | Biomechanical equipment, clinical performance measures, self-reported questionnaires, interviews, adverse events |
| Psychometrics | Clinical applicability, Validity, reliability, responsiveness, sensitivity, and specificity. |

## Results

We will narrate data and use an iterative method to summarize concepts and themes. These summaries will be presented in table format. FSR terms will be defined, and studies will be categorized based on the scoping review aims (e.g., FSR assessment), study type (e.g., cross-sectional or cohort), and population characteristics (e.g., neurological disease or orthopedic injuries).

We expect to be able to identify definitions or define the FSR transition phases and events and describe the task’s key movements and common strategies in addition to listing FSR outcome measures and reporting their psychometrics. We anticipate discussing potential motor and socio-cultural factors influencing FSR. Finally, we will demonstrate the limitations and gaps in FSR literature to inform future research directions.

## Discussion

This scoping review protocol aims to provide a comprehensive overview of the measurement properties of various FSR assessment tools and to identify the body structures and functions that influence the performance of individuals in this task. Accurate assessment and understanding of the measurement properties of existing assessments, as well as the influencing factors, can enhance the clinical assessment process. By gathering, summarizing, and synthesizing the relevant literature, this scoping review will offer valuable insights into the strengths and weaknesses of current assessment measures and help inform diagnostic clinical decision-making in the field of rehabilitation.

This review will allow healthcare professionals and researchers to make more informed choices regarding selecting and adapting FSR assessment tools. Importantly, this review may reveal gaps in the existing literature, prompting further research in areas that require more attention. The knowledge generated from this review will contribute to advancing evidence-based practice in clinical assessment and assist clinicians in targeting different FSR motor control mechanisms.

## Data Availability

All data produced in the present work are contained in the manuscript

## Notes

### Competing Interest Statement

The authors have declared no competing interest.

### Funding Statement

This study did not receive any funding

## References

Acker, S. M., Cockburn, R. A., Krevolin, J., Li, R. M., Tarabichi, S., & Wyss, U. P. (2011). Knee Kinematics of High-Flexion Activities of Daily Living Performed by Male Muslims in the Middle East. The Journal of Arthroplasty, 26(2), 319–327. 10.1016/j.arth.2010.08.003

Alomar, J. A., Catelani, M. B. C., Smith, C. N., Patterson, C. G., Artman, T. M., & Piva, S. R. (2020). Validity and Responsiveness of Floor Sitting-Rising Test in Post– Total Knee Arthroplasty: A Cohort Study. Archives of Physical Medicine and Rehabilitation, 101(8), 1338–1346. 10.1016/j.apmr.2020.03.012

Araújo, C. G. S., Castro, C. L. B., Franca, J. F. C., & Araújo, D. S. (2020). Sitting–rising test: Sex- and age-reference scores derived from 6141 adults. European Journal of Preventive Cardiology, 27(8), 888–890. 10.1177/2047487319847004

Ardali, G., Brody, L. T., States, R. A., & Godwin, E. M. (2019). Reliability and Validity of the Floor Transfer Test as a Measure of Readiness for Independent Living Among Older Adults. Journal of Geriatric Physical Therapy, 42(3), 136–147. 10.1519/JPT.0000000000000142

Borchers, E. E., McIsaac, T. L., Bazan-Wigle, J. K., Elkins, A. J., Bay, R. C., & Farley, B. G. (2019). A physical therapy decision-making tool for stratifying persons with Parkinson’s disease into community exercise classes. Neurodegenerative Disease Management, 9(6), 331–346. 10.2217/nmt-2019-0019

Brito, L. B. B. de Ricardo, D. R., Araújo, D. S. M. S. de, Ramos, P. S., Myers, J., & Araújo, C. G. S. de. (2014). Ability to sit and rise from the floor as a predictor of all-cause mortality. European Journal of Preventive Cardiology, 21(7), 892–898. 10.1177/2047487312471759

Cattuzzo, M. T., de Santana, F. S., Safons, M. P., Ré, A. H. N., Nesbitt, D. R., Santos, A. B. D., Feitoza, A. H. P., & Stodden, D. F. (2020). Assessment in the Supine-To-Stand Task and Functional Health from Youth to Old Age: A Systematic Review. International Journal of Environmental Research and Public Health, 17(16), 5794. 10.3390/ijerph17165794

Ferraz, D. D., Trippo, K. V., Duarte, G. P., Neto, M. G., Bernardes Santos, K. O., & Filho, J. O. (2018). The Effects of Functional Training, Bicycle Exercise, and Exergaming on Walking Capacity of Elderly Patients With Parkinson Disease: A Pilot Randomized Controlled Single-blinded Trial. Archives of Physical Medicine and Rehabilitation, 99(5), 826–833. 10.1016/j.apmr.2017.12.014

Fleming, J., Brayne, C., & Cambridge City over-75s Cohort (CC75C) study collaboration. (2008). Inability to get up after falling, subsequent time on floor, and summoning help: Prospective cohort study in people over 90. BMJ (Clinical Research Ed.), 337, a2227. 10.1136/bmj.a2227

Fukuichi, A., & Sugamura, G. (2022). Sitting posture and moral impression formation: A focus on traditional Japanese sitting posture (Seiza). Journal of Physical Education and Sport, 22(2), 503–511. 10.7752/jpes.2022.02063

Gurley, R. J., Lum, N., Sande, M., Lo, B., & Katz, M. H. (1996). Persons found in their homes helpless or dead. The New England Journal of Medicine, 334(26), 1710– 1716. 10.1056/NEJM199606273342606

Hewes, G. W. (1957). The Anthropology of Posture. Scientific American, 196(2), 122– 133.

Lyman, S., Omori, G., Nakamura, N., Takahashi, T., Tohyama, H., Fukui, N., Ikeda, H., Sasho, T., Saito, T., Hayashi, Y., & Deie, M. (2019). Development and validation of a culturally relevant Japanese KOOS. Journal of Orthopaedic Science: Official Journal of the Japanese Orthopaedic Association, 24(3), 514–520. 10.1016/j.jos.2018.11.014

Mohamed, C. R., Nelson, K., Wood, P., & Moss, C. (2015). Issues post-stroke for Muslim people in maintaining the practice of salat (prayer): A qualitative study. Collegian (Royal College of Nursing, Australia), 22(3), 243–249. 10.1016/j.colegn.2014.01.001

Mulholland, S. J., & Wyss, U. P. (2001). Activities of daily living in non-Western cultures: Range of motion requirements for hip and knee joint implants. International Journal of Rehabilitation Research, 24(3), 191–198.

Ng, S. S. M., Fong, S. S. M., Chan, W. L. S., Hung, B. K. Y., Chung, R. K. S., Chim, T. H. T., Kwong, P. W. H., Liu, T.-W., Tse, M. M. Y., & Chung, R. C. K. (2016). The sitting and rising test for assessing people with chronic stroke. Journal of Physical Therapy Science, 28(6), 1701–1708. 10.1589/jpts.28.1701

Quinn, L., Riley, N., Tyrell, C. M., Judd, D. L., Gill-Body, K. M., Hedman, L. D., Packel, A., Brown, D. A., Nabar, N., & Scheets, P. (2021). A Framework for Movement Analysis of Tasks: Recommendations From the Academy of Neurologic Physical Therapy’s Movement System Task Force. Physical Therapy, 101(9), pzab154. 10.1093/ptj/pzab154

Weingarten, G., & Kaplan, S. (2015). Reliability and validity of the Timed Floor To Stand Test-Natural in school-aged children. Pediatric Physical Therapy: The Official Publication of the Section on Pediatrics of the American Physical Therapy Association, 27(2), 113–118. 10.1097/PEP.0000000000000118

